# Estimation of Physiological Metrics from Resting ECGs Using Deep Learning in the UK Biobank, Including submaximal exercise derived V̇O_2_max, Body Fat Percentage, and Grip Strength

**DOI:** 10.64898/2026.05.09.26352818

**Authors:** Isabelle Mankowski, Elias Pinter, I-Min Lee, Gunnar Rätsch, David Brüggemann, Olga V. Demler

## Abstract

Maximal oxygen consumption 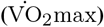 is the gold standard for cardiorespiratory fitness but requires resource-intensive physical testing. Recent reports show that machine learning models can extract additional information from ECGs, yet the potential of ECG as a source of physiological metrics remains underutilized. While routinely collected resting electrocardiograms (ECG) provide an opportunistic window into cardiorespiratory fitness, current deep learning models often struggle with cross-cohort transferability or remain dependent on active exercise data. We developed population specific models using the UK Biobank to estimate submaximal exercise derived 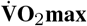(*N* = 8,540) and a panel of other physiological metrics (sample sizes up to *N* = 78,265) from resting 12-lead ECGs using Patient Contrastive Learning of Representations (PCLR), an AI based tool that converts ECG into a set of 320 features (ECG-PCLR). Data were split 80%:20% (training:test) and models were evaluated on a set-aside test subset. We demonstrate that ECG-PCLR embeddings alone can estimate submaximal 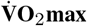 and body fat percentage with Pearson correlations (*r*) of 0.61 and 0.65, respectively. They also estimate systolic blood pressure, forced expiratory volume in 1 second (FEV1), and grip strength with *r* values from 0.31 to 0.55. Adding ECG embeddings to basic predictors (age, sex and BMI) improves submaximal 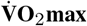 prediction by an absolute Δ*R*^2^ of 8% and by 1% to 13% for other physiologic parameters.

## Introduction

Cardiorespiratory fitness (CRF) is a vital indicator of overall health and a strong predictor of all-cause mortality^(1)^. Maximal oxygen consumption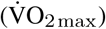, the gold-standard measure of CRF, requires costly, time-consuming Cardiopulmonary Exercise Testing^(2,3)^, limiting routine use. Deep learning on resting 12-lead electrocardiograms (ECGs) can estimate metrics invisible to human experts, including 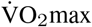 ^(4)^ and reduced left ventricular ejection fraction (LVEF)^(5)^. However, it remains unclear if these models capture intrinsic cardiac signals or merely proxy biometrics (age, sex, BMI). In this report, we explore whether 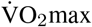 and CRF parameters can be opportunistically estimated by leveraging resting ECGs alone or with readily-available clinical characteristics. We developed and internally validated models estimating submaximal exercise-derived 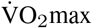, LVEF, blood pressure, body fat percentage, forced expiratory volume in 1-second (FEV1), and grip strength in the UK Biobank.

## Methods

### Study Population and Data

We included UK Biobank participants^(6)^ with resting ECGs (Field ID 20205, Instance 2, 2014-2025). Test aware submaximal exercise extrapolations of 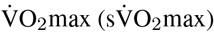 were derived from exercise ECGs at Instances 0 and 1 (2006–2010 and 2012–2013, respectively) and provided by UK Biobank (Field ID 30038) . Although not derived from direct gas exchange analysis (the gold standard), 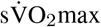 have shown moderate to strong agreement with directly measured 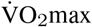 (Pearson’s *r* = 0.68 to 0.74) with no significant mean bias^(7)^, representing a validated proxy outcome. LVEF, systolic and diastolic blood pressure, body fat percentage, FEV1 and left-handed grip strength were collected at Instance 2.

### Feature Extraction and Modeling

Raw 12-lead ECG signals were preprocessed (resampled to 4096 samples, scaled to mV) and passed through a pre-trained Patient-Contrastive Learning of Representations (PCLR) ResNet encoder to extract 320-dimensional feature embeddings from raw resting ECG traces^(8)^. We compared linear (Lasso, Elastic Net) and non-linear (XGBoost, MLP) regression models.

We used three distinct modeling strategies:

- **Basic model:** Age, Sex, BMI.
- **ECG model:** 320 PCLR ECG embeddings only.
- **Full model:** Age, Sex, BMI and 320 PCLR ECG embeddings. Throughout this study, the term “ECG-only model” refers to models using pre-trained ECG-PCLR embeddings as predictors without inclusion of age, sex, or BMI.

For each outcome-specific cohort, we reserved a random 20% of participants as a held out test set and selected model hyperparameters using 10-fold crossvalidation within the remaining 80% training set. Performance was evaluated using the Coefficient of Determination (*R*^2^), Pearson correlation (*r*), and Mean Absolute Error (MAE).

## Results

### Study Population

The overall study population included all participants with an available resting ECG at Instance 2 (N = 84,125), with a mean (SD) of age: 66.2 (8.0) years and a BMI: 26.6 (4.5) kg/m^2^. The cohort was 51.9% female, predominantly white (96.2%). Mean(SD) blood pressure was 140.7 (19.4) mmHg systolic and 79.5 (10.2) mmHg diastolic. In this cohort, the mean (SD) of target physiological metrics at Instance 2 were 59.4% (6.4) for LVEF, 31.2% (8.2) for body fat percentage, 2.7 L (0.8) for forced expiratory volume in 1 second (FEV1), and 27.8 kg (10.6) for left hand grip strength. For the primary outcome, the mean estimated 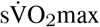 in this cohort was 29.8 (6.4) ml/kg/min at Instance

0 and 29.6 (6.7) ml/kg/min at Instance 1.

### Models of 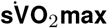 and Feature Contribution

The addition of ECG embeddings enhanced fitness estimation. While the biometric-only Basic model established a strong baseline (*R*^2^ = 0.49), the Full model achieved an *R*^2^ of 0.57. Lasso model explained over 50% of variability (*R*^2^=0.57, MAE=3.31 ml/kg/min), outperforming non-linear models such as the Multilayer Perceptron (*R*^2^=0.48, MAE=3.67 ml/kg/min). This superiority aligns with prior studies demonstrating effective linear probe transfer^(8)^, as well as prior work applying PCLR embeddings to estimate peak oxygen consumption^(4)^. The ECG-only Model achieved an *r* of 0.61 and *R*^2^=0.37, MAE=4.24 ml/kg/min) (Table 1).

**Table 1.** Incremental Value of ECG Embeddings across Physiological Metrics. Comparison of the Basic model (Age, Sex, BMI), the Full model (Basic + ECG derived embeddings) and ECG model (ECG derived embeddings only) on the held out test set. Results are reported for the Lasso regressor.

| Physiological Metric | N (test set) | Basic Model<br>(Age + Sex + BMI) |  |  | Full Model<br>(Basic + ECG) |  |  | ECG Model<br>(ECG only) |  |  |
| --- | --- | --- | --- | --- | --- | --- | --- | --- | --- | --- |
| | | $R^2$ | Pearson $r$ | MAE | $R^2$ | Pearson $r$ | MAE | $R^2$ | Pearson $r$ | MAE |
| *Primary Outcome* |  |  |  |  |  |  |  |  |  |  |
| sVO<sub>2</sub>max (Inst. 1) (ml/kg/min) | 1,708 | 0.49 | 0.70 | 3.63 | **0.57** | **0.76** | **3.31** | 0.37 | 0.61 | 4.24 |
| *Secondary Cardiovascular Metrics* |  |  |  |  |  |  |  |  |  |  |
| sVO<sub>2</sub>max (Inst. 0) (ml/kg/min) | 2,261 | 0.49 | 0.70 | 3.48 | **0.55** | **0.74** | **3.28** | 0.36 | 0.60 | 3.98 |
| LVEF (%) | 933 | 0.05 | 0.24 | 5.17 | **0.18** | **0.44** | **4.95** | 0.09 | 0.31 | 4.82 |
| Systolic BP (mmHg) | 12,996 | 0.16 | 0.40 | 14.21 | **0.26** | **0.51** | **13.20** | 0.18 | 0.43 | 13.83 |
| Diastolic BP (mmHg) | 12,997 | 0.07 | 0.26 | 7.82 | **0.15** | **0.39** | **7.42** | 0.13 | 0.36 | 7.55 |
| *Body Composition & Physical Function* |  |  |  |  |  |  |  |  |  |  |
| Body Fat % | 9,716 | 0.81 | 0.90 | 2.80 | **0.82** | **0.91** | **2.71** | 0.42 | 0.65 | 4.93 |
| FEV1 (L) <sup>1</sup> | 7,459 | 0.51 | 0.71 | 0.40 | **0.52** | **0.72** | **0.39** | 0.28 | 0.53 | 0.50 |
| Grip Strength (kg) | 15,653 | 0.51 | 0.72 | 5.71 | **0.52** | **0.72** | **5.65** | 0.30 | 0.54 | 6.99 |

Data for all parameters, unless indicated otherwise, were collected at instance 2. sVO<sub>2</sub>max is derived from submaximal exercise.
<sup>1</sup> Forced expiratory volume in 1 second (FEV1) measured with spirometry.

We investigated whether the Full model used ECG features as proxies of basic clinical characteristics (age, sex and BMI) or captured unique physiological signals. In the Basic model, biological sex (52.6%) and BMI (29.7%) were the dominant contributors. However, in the Full model, the PCLR embeddings collectively accounted for 61.0% of the contribution, relegating sex (21.2%) and BMI (10.9%) to secondary roles. The coefficient weighted contribution analysis (Figure 1a) suggests that ECG derived embeddings contributed substantially to the fitted model, but this analysis should not be interpreted as proof that the embeddings capture intrinsic cardiac physiology independent of demographic or anthropometric correlates.

**Fig. 1.**
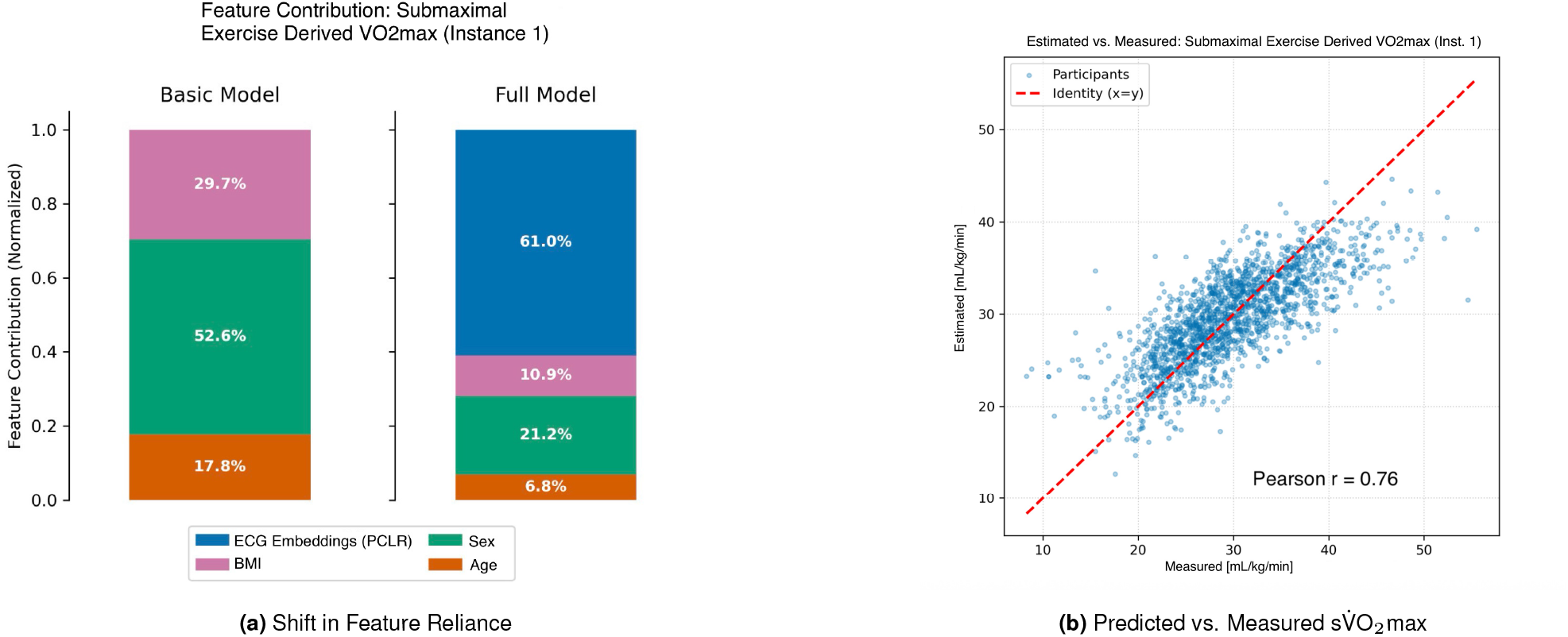
Lasso Model Feature Importance and Performance. **(a)** Comparison of aggregated feature contributions for submaximal exercise derived 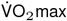 prediction. Consistent with prior literature, the Basic model relies heavily on biological sex (52.6%) and BMI (29.7%), while the Full model shifts 61.0% of predictive weight to PCLR embeddings (ECG signals). Feature contributions are calculated as the mean absolute product of standardized input features and their corresponding coefficients (|*β · X*_std_ |). To determine categorical importance, these values are cumulatively summed within each group and normalized to sum to 100%. While these embeddings contribute substantially to the model’s performance, this does not definitively confirm that they isolate intrinsic cardiac physiology apart from age, sex and BMI. **(b)** Scatter plot of predicted vs. measured 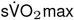 showing alignment with the identity line, despite regression to the mean at extreme fitness deciles. 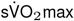 estimated from submaximal exercise testing.

The Full model exhibited regression to the mean, over-predicting low fitness (<20 ml/kg/min) and under-predicting high-performing individuals (>45 ml/kg/min) (Figure 1b).

### Physiological Versatility: models of body composition, cardiovascular metrics, and physical function

The ECG-only model demonstrated standalone predictive power. Although models of body composition metrics heavily relied on BMI, estimates from ECG-only model achieved a strong Pearson correlation with body fat percentage (*r*=0.65), suggesting that resting ECGs contain measurable information correlated with body composition. The ECG-only model estimated 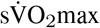, FEV1, and grip strength with *r*=0.61, 0.53 and 0.54 respectively. Addition of ECG-PCLR embeddings to Basic model notably improved cardiac-specific metrics such as LVEF (*r*=0.24 vs *r*=0.44) and systolic blood pressure (*r*=0.40 vs *r*=0.51), though achieved predictive LVEF performance remained modest (Table 1).

## Conclusions

The Full model explained more than half of the variability in 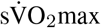, FEV1, and grip strength in the held-out test subset. ECG-only models showed strongest performance for body fat percentage (*r* = 0.65), followed by 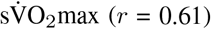 and grip strength (*r* = 0.54). The addition of ECGs-derived PCLR embeddings to age, sex, and BMI improved prediction across multiple cardiopulmonary and physiological metrics, suggesting that resting ECGs encode incremental physiological information beyond standard demographic and anthropometric characteristics. While coefficient-weighted contribution analysis indicated substantial contribution of ECG embeddings to the Full model, future studies are needed to clarify the extent to which these representations reflect intrinsic cardiac physiology versus correlated demographic or anthropometric structure.

Unlike approaches requiring subjective questionnaires or multi-day wearable monitoring^(9,10)^, we used framework that derives physiological estimates from a standard 10-second resting ECG using fixed pre-trained deep learning embeddings. Khurshid et al. ^(4)^ previously demonstrated that PCLR-derived ECG embeddings combined with penalized regression can estimate CPET-measured peak oxygen consumption in clinical cohorts. We extend this paradigm to a large population-based UK Biobank cohort and to multiple physiological traits including 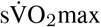 and seven additional physiological traits.

This study has several notable strengths and limitations. First, the 1 to 13 year temporal gap between resting ECGs and 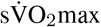 can introduce potential label mismatch and signal attenuation due to aging, biological changes, and treatments.

Our Full model achieved an *R*^2^ of 0.57 for Instance 1, while performance was slightly lower for a longer gap for Instance 0 (*R*^2^ = 0.55). We hypothesize that a shorter temporal gap between ECG acquisition and fitness measurement would reduce expected attenuation from label temporal mismatch, potentially yielding even stronger estimations of cardiorespiratory capacity. The PCLR encoder was trained on *>* 3*M* ECGs in external data^(8)^ and showed robust performance in this cohort. These findings support the utility of contrastive learning approaches, and PCLR embeddings specifically, for representation learning from resting ECGs. Evaluating further external transportability and generalization for these ap-proaches in diverse datasets remains crucial for future work. Finally, although the UK Biobank’s nonclinical recruitment likely attenuated confounding by indication, relative to hospital cohorts, healthy volunteer bias and limited ancestry diversity remain.

In summary, pre-trained PCLR embeddings derived from resting 12-lead ECGs captured information associated with multiple dimensions of cardiorespiratory fitness and physiological function. Because resting ECGs are routinely acquired in clinical practice, these findings support further investigation of ECG foundation-model embeddings as scalable tools for passive physiological phenotyping and opportunistic fitness assessment, subject to external validation and deployment considerations.

## Data Availability

The data used in this study are available from the UK Biobank via standard access procedures.

## ETHICS APPROVAL AND CONSENT

Ethics approval for this study was obtained from the Cantonal Ethics Committee of Zurich and Institutional Review Board at Brigham and Women’s Hospital.

## AUTHOR CONTRIBUTIONS

All authors contributed to the conception and design of the study. Material preparation: IM, OD, DB data collection: IM, OD, DB analysis: IM, OD. All authors read and approved the final manuscript.

## DATA AVAILABILITY

This research was conducted using the UK Biobank Resource under Application Number 206575. We thank the UK Biobank and UK Biobank participants for providing the data resource. The data used in this study are available from the UK Biobank via standard access procedures.

## CODE AVAILABILITY

The code used for ECG preprocessing (resampling and scaling) and the model training scripts are available at https://github.com/CodeBellee/Deep-Learning-Fitness-Estimation-ECG.git.

